# Development and performance of a population-based risk stratification model for COVID-19

**DOI:** 10.1101/2021.05.25.21257783

**Authors:** Emili Vela, Gerard Carot-Sans, Montse Clèries, David Monterde, Xènia Acebes, Adrià Comella, Luís García Eroles, Marc Coca, Damià Valero-Bover, Jordi Piera-Jiménez, Pol Pérez Sust

## Abstract

The shortage of recently approved vaccines against the severe acute respiratory syndrome coronavirus (SARS-CoV-2) has highlighted the need for evidence-based tools to prioritize healthcare resources for people at higher risk of severe coronavirus disease 2019 (COVID-19). Current evidence indicates that age is far from accurate in identifying the risk of severe illness; furthermore, the count of individual risk factors has limited applicability to population-based “stratify-and-shield” strategies. We developed a COVID-19 risk stratification system that allows allocating people into four mutually-exclusive risk categories based on multivariate models for hospital admissions, transfer to intensive care unit (ICU), and mortality among the general population. The model was developed using clinical, hospital, and epidemiological data from the entire population of Catalonia (North-East Spain; 7.5 million people) and validated using an independent dataset of 218,329 individuals with PCR-confirmed COVID-19, who were infected after developing the model. This showed high discrimination capacity, with an area under the curve of the receiving operating characteristics of 0.85 (95% CI 0.85 – 0.85) for hospital admissions, 0.86 (0.86 – 0.97) for ICU transfers, and 0.96 (0.96 – 0.96) for deaths. Our results provide clinicians and policymakers with an evidence-based tool for prioritizing COVID-19 healthcare resources other population groups aside from those with higher exposure to SARS-CoV-2 and frontline workers.

## Introduction

The recently approved vaccine against the severe acute respiratory syndrome coronavirus (SARS-CoV-2) is an expected game-changer of the COVID-19 pandemic. However, the massive number of doses needed to achieve herd immunity will likely lead to a scarcity of the marketed vaccines. This scenario, which may worsen if long-term immunity is not achieved [1], will force governments to establish priority criteria for accessing vaccines. This prioritization also applies to other healthcare resources needed for preventive strategies such as screening campaigns, awareness programs, and early administration of specific therapies that are not widely available.

Aside from protecting highly exposed individuals like healthcare workers, the risk of serious illness seems to be the most reasonable criterion to prioritize access to COVID-19 resources based on a “stratify-and-shield” strategy [2]. In the absence of a consensus framework for COVID-19 risk allocation, age at the cutoff of 65 years has been proposed as a criterion for targeting populations for vaccine prioritization [3]. However, evidence indicates that age is far from accurate in identifying the risk of severe illness [4], and the idea of using age as the sole criterion for prioritization has raised ethical concerns [5]. To date, various risk factors associated with the severe illness have been identified, including clinical, demographic, and socioeconomic characteristics [6–8]. Based on these factors, various prediction models for COVID-19 have been proposed, most of them based on data from cohorts of limited size or aimed at estimating risk in specific populations like hospitalized patients [9]. An exception to this trend is the risk index developed by DeCapprio et al., which used a nationwide approach to develop a numeral index for predicting the risk of complications due to upper respiratory infections among the general population [10]. Alternatively, we propose a model that allows classifying the general population into mutually exclusive risk groups for severe COVID-19. In countries with centralized electronic health records, this model may help policymakers in resource prioritization and planning (e.g., vaccines, diagnostic tests, and hospital and intensive care unit [ICU] beds) and clinicians in making therapeutic decisions.

Using whole-population data on hospitalizations, ICU transfers, and deaths due to COVID-19 in our area, we developed a population-based model intended to group people according to their risk of serious events due to COVID-19. Based on the ideal characteristics of such stratification system suggested elsewhere [4], we sought a system that was population-based (i.e., all individuals in a community could be assigned to mutually-exclusive groups), accessible (i.e., it must be based on information available and accessible to all healthcare professionals), understandable (i.e., it must be easily explained to policymakers and citizens), discriminatory (i.e., individuals could be allocated in a discrete list of strata), and suitable for local implementation.

## Methods

### Data sources

We retrospectively retrieved data from administrative databases in Catalonia, a North-East region in Spain with a population of 7.5 million people. Data on potential predictors were retrieved from the Catalan Health Surveillance System (CHSS), which systematically collects data regarding diagnoses, individual income, and resource utilization from both hospital and primary care settings [11]. Data on outcomes associated with SARS-CoV-2 infection were retrieved from the epidemiological surveillance system in the SARS-CoV-2 registry (RSACovid-19) [12,13]. The stratification model was built using data collected between March 1 and September 15, 2020 (development period), which encompassed the first wave of the COVID-19 outbreak in our area and a period between waves. Data for model validation had been collected between September 16 and December 27, 2020 (i.e., the date the first vaccine was administered in Catalonia) (validation period).

All data were handled according to the General Data Protection Regulation 2016/679 on data protection and privacy for all individuals within the European Union and the local regulatory framework regarding data protection. Data from different health administrative databases were linked and de-identified by a team not involved in the study analysis; study investigators only had access to a fully anonymized database. The retrospective use of healthcare data was approved by the Independent Ethics Committee of the IDIAP Jordi Gol (Spain), which waived the need for obtaining informed consent for data utilization.

### Predictors

We considered all variables stored in the CHSS database, including demographic data (i.e., age and sex), resource utilization (e.g., admission to nursing homes), lifestyle information (e.g., smoking, and alcohol abuse), current and past diagnoses (including psychiatric disorders), and socioeconomic status. The global comorbidity burden (or patient complexity) was stratified using the adjusted morbidity groups (GMA, *Grups de Morbiditat Ajustada*), a population-based tool for health-risk assessment [14,15]. The GMA tool considers the weighted sum of all chronic conditions, the number of systems affected, and acute diagnoses present at the time that may increase patient complexity. Individuals are grouped into four health-risk categories defined using the risk distribution of the entire population: (1) baseline risk (healthy stage, including GMA scores up to the 50^th^ percentile of the total population), (2) low risk, 50^th^ to 80^th^ percentiles, (3) moderate risk, 80^th^ to 95^th^ percentiles, and (4) high risk, above the 95^th^ percentile. Socioeconomic status was stratified according to pharmaceutical co-payment groups, which are based on annual income, as follows: very low (i.e., recipient of rescue aid measures), low (i.e., less than € 18,000), middle (i.e., € 18,000 to € 100,000), and high (i.e., >€ 100,000).

### Outcomes

We analysed three outcomes associated with severe COVID-19: hospital admission, transfer to intensive care unit (ICU), and death. The scarcity of PCR tests during the pandemic precluded the testing of all suspected cases of COVID-19. For that reason, we considered the COVID-19 diagnosis according to either molecular criteria (positive result with a PCR or serological test) or clinical/epidemiological criteria, as officially established by the RSACovid-19. Owing to the shortage of ICU beds during the first wave (March 03 to July 15, 2020), the start of invasive mechanical ventilation was considered an ICU transfer, irrespective of an ICU admission registry. All deaths related to COVID-19 were included, whether they had been hospitalized or not.

### Statistics

The dataset for developing the stratification model included all individuals with any of the investigated outcomes within the development period, irrespective of the time of COVID-19 diagnosis. We used generalized linear models (Poisson regression) to build multivariate models for hospitalizations, ICU transfers, and deaths due to COVID-19. The models were created using a “stepwise-forward” approach based on the Akaike Information Criterion (AIC), in which a naïve model is sequentially complemented with the most relevant variables, eventually leading to the main effects model. The models also included all significant first-order interactions between selected variables and sex. Owing to its non-linear behaviour, age was introduced into the model as a continuous variable plus an additional quadratic term. The models provided individual-level estimates of the probability for each outcome (i.e., hospitalization, transfer to ICU, and death) for the entire population of Catalonia. The accuracy of the three models was assessed using the area under the curve of the receiving operating characteristics (AUC ROC). The four risk strata were defined by crosslinking the three categorized probabilities.

The stratification model was validated using an independent dataset of all individuals with a positive PCR result for SARS-CoV-2 infection in a respiratory specimen within the validation period. The goodness of fit of the model was assessed using the AUC ROC and the corresponding 95% confidence interval for each outcome. All analyses were performed using R statistical software, version R-4.0.0.

## Results

### Model development

The generalized linear models were built from data on 41,468 hospitalizations, 7,987 ICU transfers, and 15,262 deaths (all of them associated with COVID-19), which occurred during the first six months of the outbreak in Catalonia (development period). The variables included in each model, their contribution to the model, and the results of the AUC ROC analysis are summarized in Figure S1 (Supplementary appendix). The crosslinking probabilities of the three outcomes resulted in four mutually exclusive groups at low, moderate, high, and very high risk. Figure 1 shows the proportion of individuals allocated to each group and the age distribution across risk groups for the reference population. Model calibration showed low discrepancy between observed and expected cases during the development period (Figure S2). ROC AUC (95% CI) for hospitalizations, ICU transfers, and deaths were 0.82 (0.814 – 0.82), 0.83 (0.83 – 0.84), and 0.96 (0.96 – 0.96), respectively.

**Fig 1.**
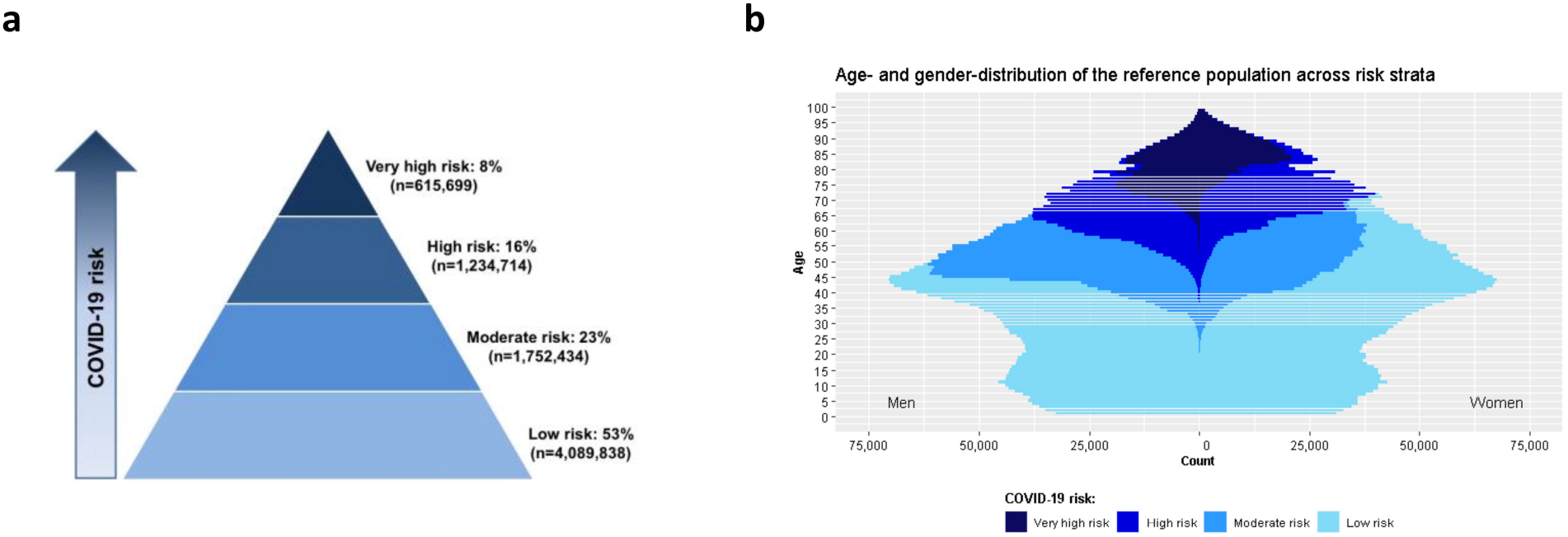
Distribution of the reference population (i.e., Catalonia, 7,697,069 inhabitants) across risk groups. **a**: percentage of individuals allocated in each risk group. **b**: age distribution across risk groups

Figure S3 summarizes the demographic and clinical profile of individuals allocated in each risk group. Briefly, the low-risk group had 55% women, with a median age of 26 years (IQR 13 – 38) and a very low prevalence of comorbidities; this group covered the healthy population. Individuals in the moderate-risk group were mostly men (66%) with a median age of 50 years (IQR 45 – 55) and a low comorbidity burden. A remarkable percentage of individuals diagnosed with AIDS (43.4%) or severe psychiatric disorders (30.7%) among the overall population fell into this group. The high-risk group had 51% of women with median age of 67 years (IQR 62 – 73). This group typically included middle-aged adults with cardiovascular risk factors; 54.6% of all individuals with hypertension, 43.5% of those with hyperlipidemia, and 35.6% of those with obesity fell into this group. The very high-risk group had 45% women with a median age of 82 years (IQR 76 – 87). This group included almost all people institutionalized in a nursing home (91.6%), diagnosed with dementia (89.7%), and receiving domiciliary care (87.6%). A remarkable percentage of individuals with kidney failure (64.8%), heart failure (69.5%), ischemic heart disease (53.8%), and stroke (51.6%) among the overall population also fell into the very high-risk group.

### Validation of the stratification model

The weekly rate of hospitalizations among the general population increased with risk groups during the entire period, being the differences between groups more pervasive during waves (Figure 2A). The other two outcomes also displayed an increasing trend across risk groups. However, the rate of ICU transfers was similar in the very high- and high-risk groups during the second wave, and mortality clearly stood out among the individuals of the very high-risk group during the two waves (Figure 2B and 2C, respectively).

**Fig 2.**
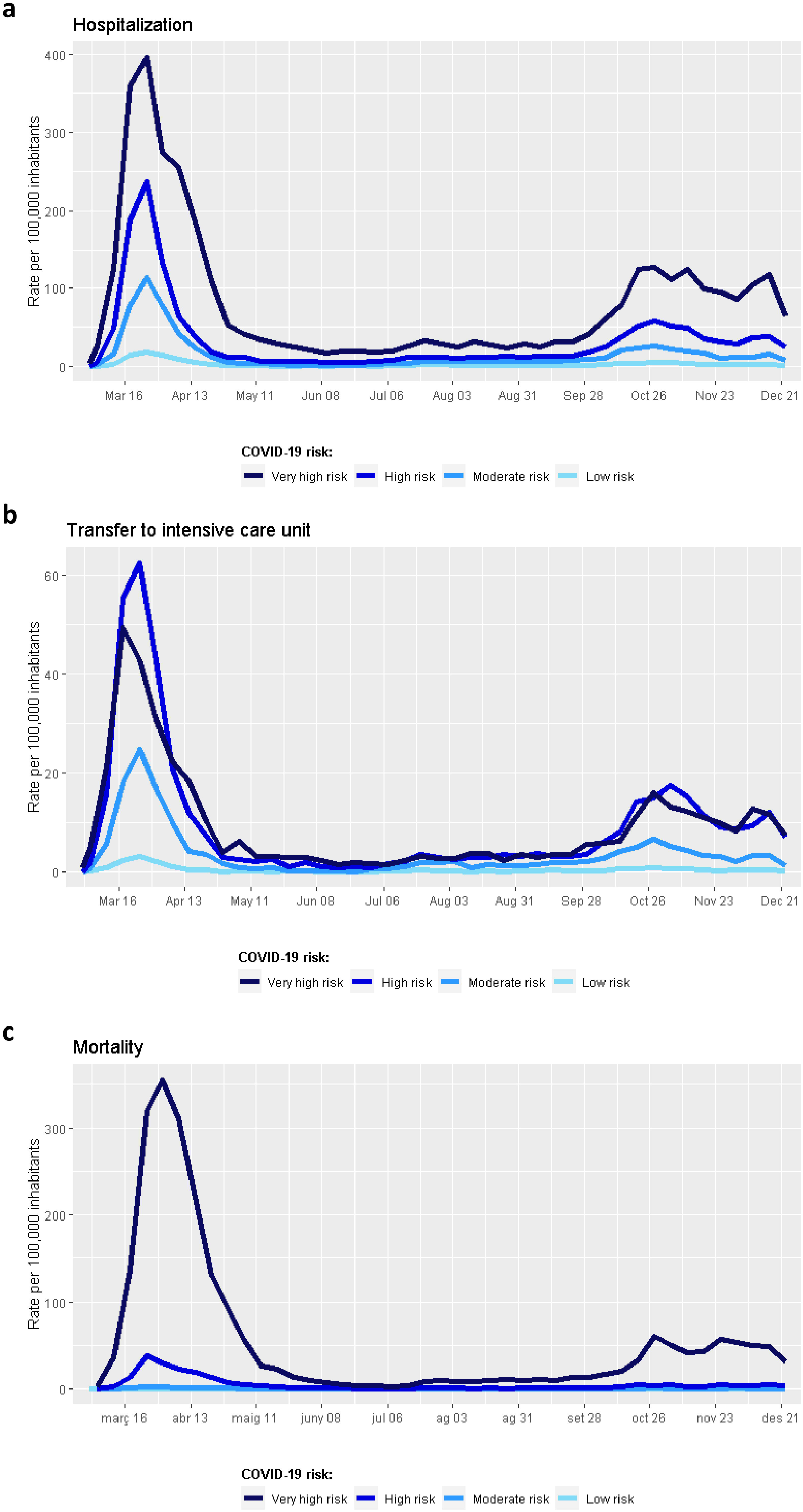
Longitudinal analysis of outcome rate within the first 10 months of the COVID-19 pandemic in Catalonia. Results are presented as the incidence rate at the population level and stratified according to COVID-19 risk group. **a**: Hospital admissions due to COVID-19. B: Transfer to an intensive care unit (ICU) due to COVID-19. **b**: Death due to COVID-19

The independent dataset for model validation included 218,329 individuals with PCR-confirmed COVID-19 diagnosis. Of these, 17,235 were admitted to hospital during the validation period, 3,450 were transferred to the ICU, and 3,852 died. Figure 3 shows the incidence rate of each outcome among individuals infected during the validation period. Hospitalization rate among infected individuals progressively increased across risk groups (Figure 3A). The rate of ICU transfer was higher in the high-risk group than the very high-risk group (Figure 3B). Lethality progressively increased from the low-risk to the high-risk group and sharply increased in the very high-risk group (Figure 3C). AUC ROC (95% CI) for hospitalization, ICU transfer, and death within the validation period was 0.85 (0.85 – 0.85), 0.86 (0.86 – 0.97), and 0.96 (0.96 – 0.96), respectively.

**Fig 3.**
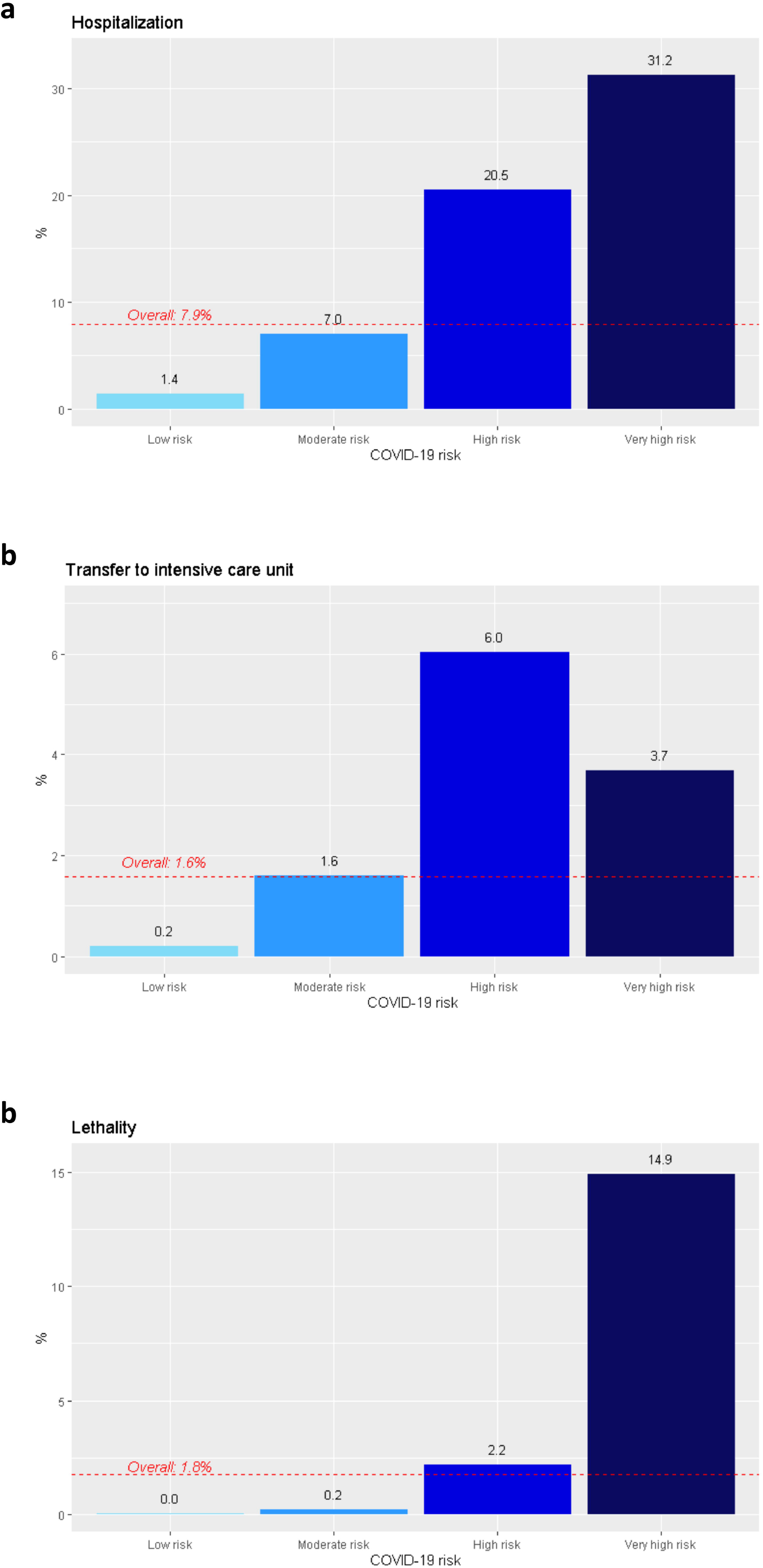
Proportion of individuals with PCR-confirmed COVID-19 (N= 218,329) who experienced each of the events within the validation period (from September 16 to December 27, 2020). The dotted red line shows the overall event rate. **a:** Hospital admission (n=17,235). **b:** Transfer to an intensive care unit (ICU) (n=3,450). **c:** Lethality (n=3,852)

## Discussion

We designed a population-based risk model aimed at stratifying the general population into mutually exclusive groups at risk of COVID-19 severe illness or death. The model showed adequate goodness of fit for hospital admissions, ICU transfers, and death. When tested on an independent dataset of PCR-confirmed COVID-19 individuals, the stratification model showed high discrimination capacity for the three outcomes. The highest differences between risk groups were observed for hospitalization rate. The frequency of ICU transfer was higher in the high-risk group than in the very high-risk group, probably because older and more frail people, typically in the very high-risk group, are often excluded from invasive practices to prevent therapeutic obstinacy. The mortality rate was notably higher in the very high-risk group than in other risk groups.

The living systematic review of the COVID-19 precise consortium identified 107 prognostic models for patients with COVID-19 diagnosis [9]. However, most of these models target individuals admitted to hospital or presenting at general practitioner with symptoms suspicious of COVID-19. Alternatively, we used data from the general population to develop a model that could provide a risk estimate, irrespective of the disease stage. This feature is important for prioritizing interventions and resources for people at risk of more severe outcomes in the event of SARS-CoV-2 infection. Following a similar population-based approach, DeCapprio et al. modelled nationwide data to develop an index to predict complications due to upper respiratory infections (as a proxy for vulnerability to COVID-19) among the general population [10]. The model showed a AUROC 0.81, close to that found in our analysis using nationwide data from COVID-19 patients (i.e., 0.85, 0.86, and 0.96 for hospital admission, ICU transfer, and death, respectively). Besides the COVID-19 specificity, our model is novel in providing a stratification system that allows allocating the general population into mutually exclusive risk groups, irrespective of the presence of symptoms. This approach offers policymakers of countries with centralized healthcare databases a helpful tool for prioritizing resources under a “stratify-and-shield” strategy. The discrimination capacity of our model when applied to an independent dataset of PCR-confirmed COVID-19 patients infected after the development period indicates that the model is also suitable for supporting therapeutic decision-making when managing COVID-19 cases.

In line with previous (i.e., early and recent) analysis of COVID-19 risk [7,8,16,17], we found that age and underlying conditions such as diabetes, arterial hypertension, and cardiovascular diseases significantly contributed to the risk of severe disease outcomes, particularly hospital admission. However, rather than individual diagnoses, the comorbidity burden was a stronger predictor of hospital admissions and deaths. Of note, unlike variables such as age or a particular diagnosis, which are homogeneous across countries and studies, the measurement of the comorbidity burden is challenged by the lack of consensus for defining and weighting chronic conditions to be considered [18,19]. The GMA stratification tool used in our model has shown high accuracy in predicting the use of healthcare services, including hospital admission [20,21]. In this regard, the inclusion of the comorbidity using other multimorbidity measures might change the performance of the model.

Our analysis was strengthened by the consistent performance of the model in two different periods and independent datasets. We deemed the analysis covering the two waves important because the overburdening of the healthcare system and resource shortage experienced during the first wave (i.e., when the model was developed) might act as important confounders. Another strength was the possibility of using data from the entire population. On the other hand, the use of administrative databases of data collected in routine care may limit the inclusion of all variables with potential influence on severe illness or death. Some of these variables (e.g., inflammatory biomarkers on admission, associated with poorer outcome [22]) were clearly inadequate for a risk model aimed at stratifying the entire population. However, other variables such as the blood group, potentially involved in COVID-19-associated respiratory failure [23], is not routinely included in the source databases of our analysis and could not be added to the model. This limitation is common among many prediction models proposed for COVID-19 [9].

In summary, the proposed risk stratification model for COVID-19 provides policymakers with evidence-based criteria for prioritizing limited COVID-19 resources, including vaccines, treatments, and tests for preventive screening of the general population. This model can also be used for needs planning (e.g., hospital and ICU beds) and to support clinicians with dynamic risk assessment of newly diagnosed COVID-19 patients. Of note, when prioritizing healthcare resources, other criteria aside from health risk shall be considered, including high exposure to SARS-CoV-2 and the development strategic actions for pandemic containment.

## Data Availability

The datasets generated and/or analysed during the current study are not publicly accessible but are available from the corresponding author upon reasonable request.

## Declarations

### Code availability

Not applicable

### Ethics approval

The study protocol was approved by the Independent Ethics Committee of the IDIAP Jordi Gol (Spain), which waived the need for written informed consent (21/043-PCV).

### Authors’ contributions

EV, MC, and DM were responsible for the study conception and design and conducted the data analysis. EV, LG-E, XA, AC, and PP-S contributed to data collection; EV, MC, DM, GC-S, MCo, DV, and JP-J contributed to data interpretation. The manuscript was first drafted by EV and GC-S; MC, DM, MCo, DV, XA, AC, JP-J, LG-E, and PP-S revised the manuscript for significant intellectual contribution. All authors have read approved the final version of the manuscript.

