## Supplementary Appendix for "Development and performance of a population-based risk stratification model for COVID-19"

Supplementary file 1

**Figure S1.** Results of the multivariate models (Poisson regression) for explaining hospital admissions (**A**), ICU transfer (**B**), and death (**C**) due to COVID-19. Models were built using data from the entire catchment population between the development period: from March 01 to September 15, 2020.

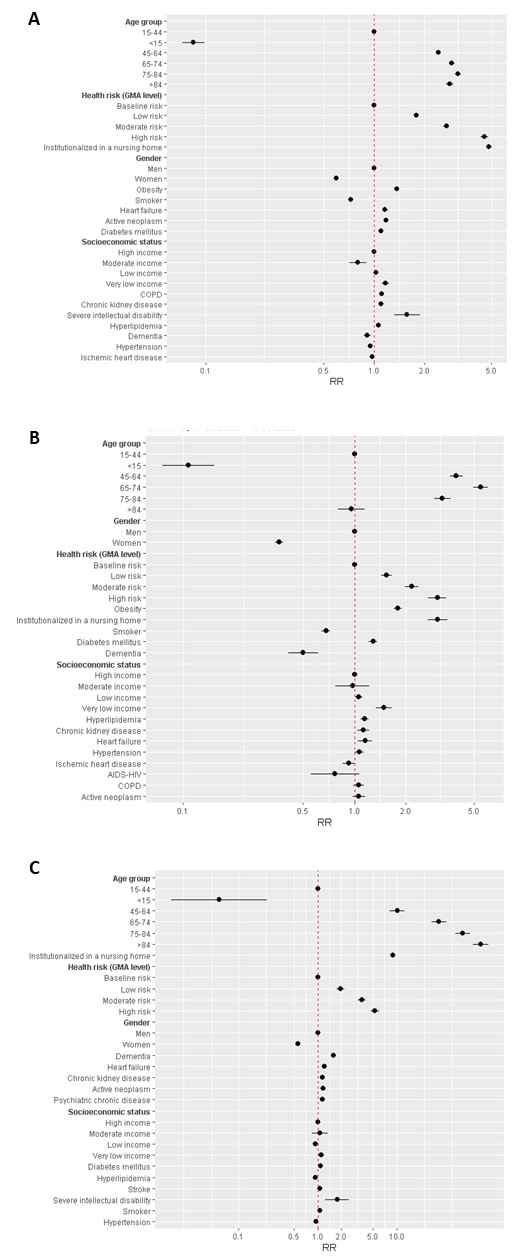

**Figure S2.** Main demographic and clinical characteristics of individuals included in each risk group: very high risk (A, B), high risk (C, D), moderate risk (E, F), and low risk (G, H).

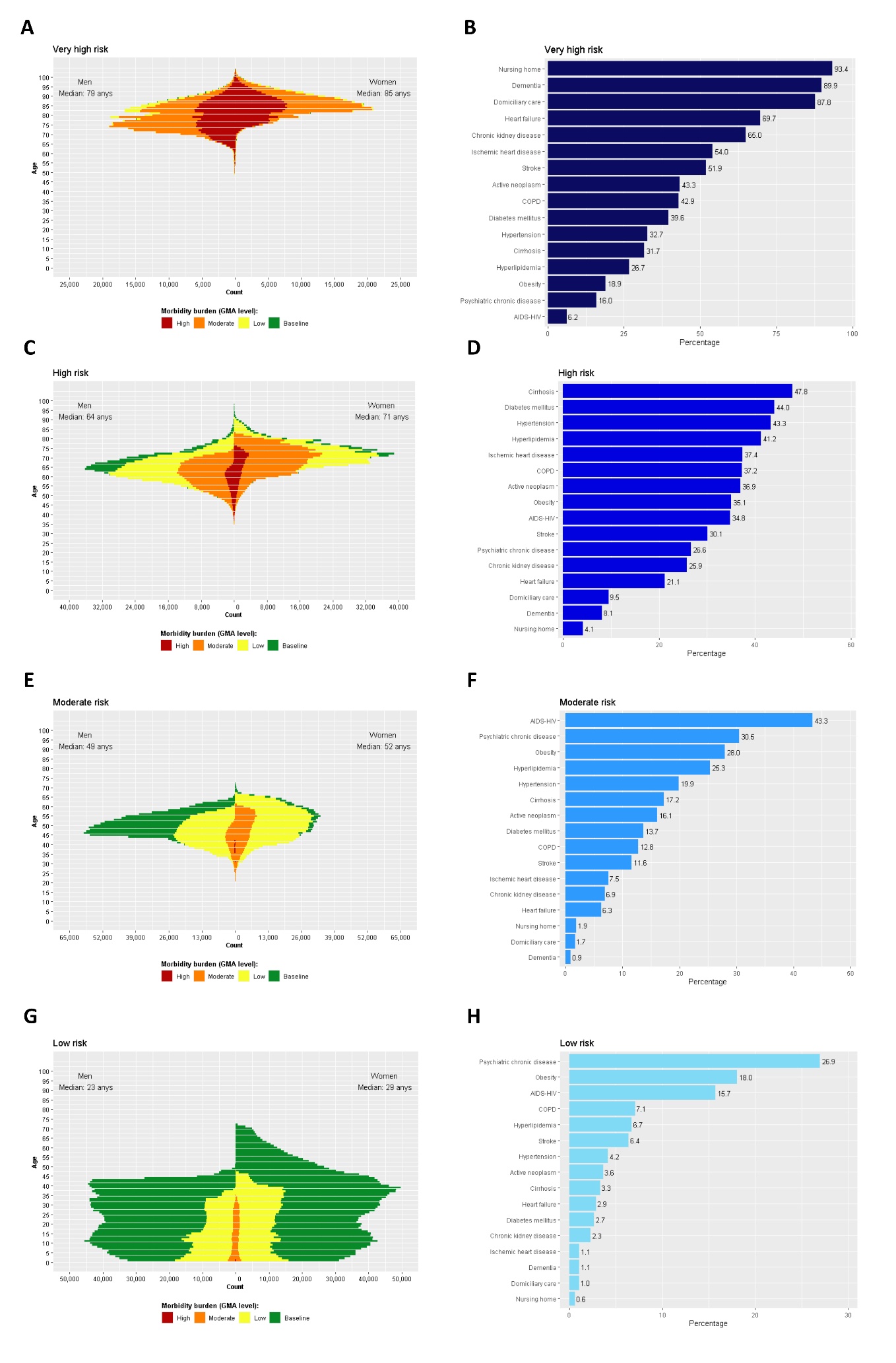

**Figure S3.** Calibration between expected and observed cases of the stratification model when considering data gathered during the development period (i.e., March 01 to September 15, 2020). Results are presented as No. of individuals experiencing the following outcomes due to COVID-19: hospital admissions (**A**), ICU transfer (**B**), and death (**C**).

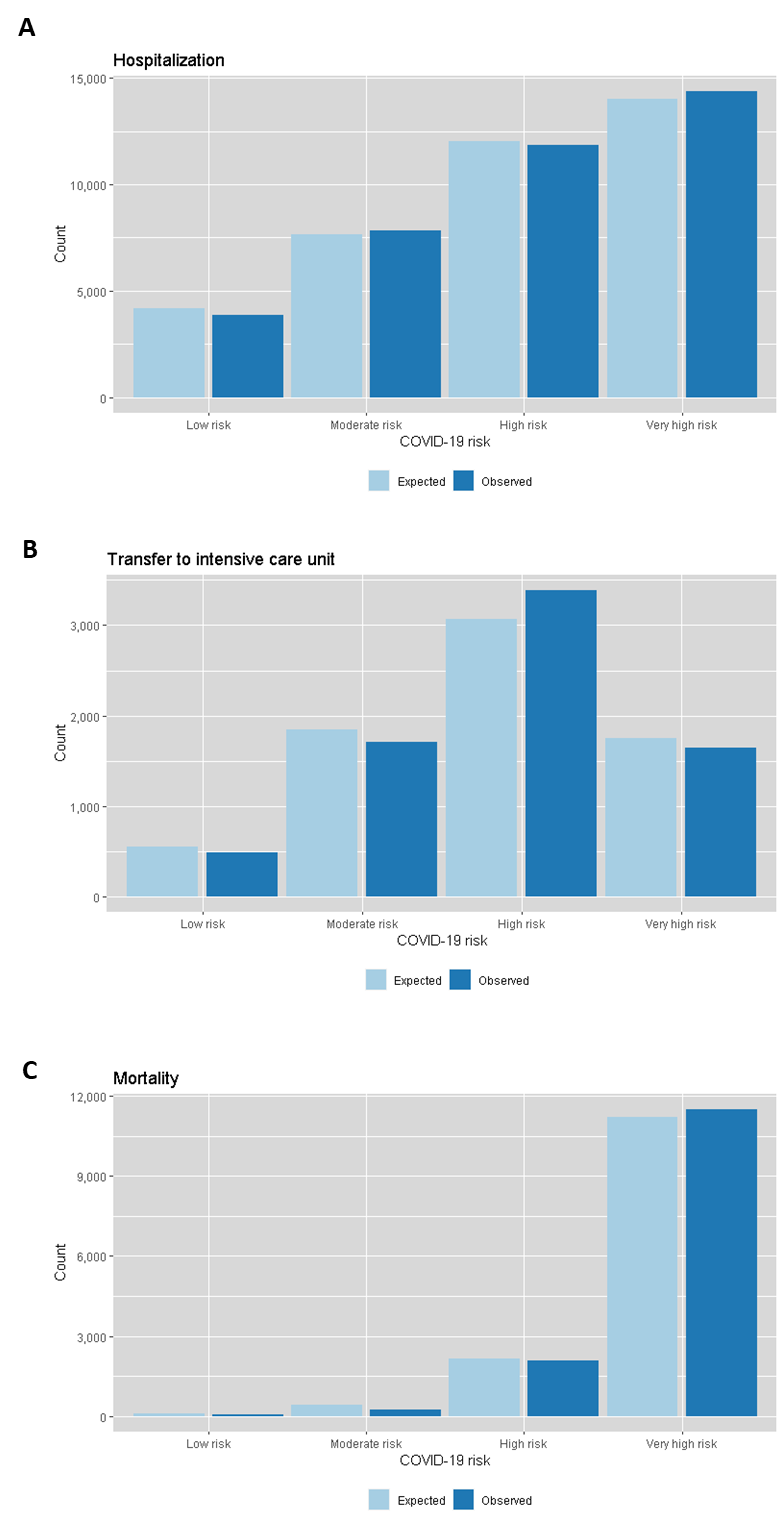
